# allofus: An R package to facilitate use of the All of Us Researcher Workbench

**DOI:** 10.1101/2024.04.10.24305611

**Authors:** Louisa H. Smith, Robert Cavanaugh

## Abstract

**Objective:** We aimed to increase research and training capacity for the All of Us community through an R package designed to reduce barriers to entry to the Researcher Workbench.

**Materials and Methods:** We developed the open-source R package `allofus`, available on the R package repository CRAN. The package provides functions that address common challenges we encountered while working with All of Us Research Program data. We tested the package with standard R unit tests and in real research projects.

**Results:** We describe how the package functions allow for an efficient workflow. We demonstrate the package’s utility by creating a cohort of All of Us participants with one year of electronic health record data prior to survey completion and no previous diagnosis of Type 2 diabetes.

**Discussion:** Despite the program’s easy-to-use tools like the Cohort Builder, using All of Us data for complex research questions requires a relatively high level of technical expertise. We developed an initial set of functions that solve problems we experienced with our own research and in mentoring student projects. In conjunction with the tutorials provided with the package, these tools can reduce the barrier for entry into the All of Us research community. The package will continue to grow and develop with the All of Us Research Program.

**Conclusion:** The `allofus` R package can help build community research capacity by increasing access to the All of Us Research Program data, the efficiency of its use, and the rigor and reproducibility of the resulting research.

## BACKGROUND AND SIGNIFICANCE

The All of Us Research Program is a groundbreaking initiative by the National Institutes of Health (NIH) to advance precision medicine using data from participants who reflect the diversity of the United States.^1^ The program collects a wide variety of data, including health and lifestyle surveys, electronic health records (EHR), genetics, wearable, and physical measurement data. These cross-cutting data sources allow researchers to explore biological, clinical, social, and environmental determinants of health and advance medical research on a personalized basis.

The data are linked and transformed into the Observational Medical Outcomes Partnership Common Data Model (OMOP CDM), an open community data standard developed by the Observational Health Data Sciences and Informatics (OHDSI) community.^2^ The OMOP CDM is designed to harmonize observational health data for efficient analyses and reliable real-world evidence.^3^ The OMOP CDM allows for the transformation of disparate databases (e.g., data from many different hospital systems) and data types into a standard format, facilitating synthesis across data sources and creating the potential for network studies across many global OMOP CDM databases. Critically, the *All of Us* program data are widely available, making the *All of Us* research program data one of the most important research assets today.

After undergoing privacy protections, the transformed database, called the Curated Data Repository (CDR), is made available to researchers on the All of Us Researcher Workbench. Here, researchers can create collaborative workspaces, which hold data and analytical scripts and provide access to compute environments. Data are stored in a Google BigQuery relational database hosted on the Workbench, accessible using SQL (structured query language), the statistical programming language R,^4^ or the general-purpose language Python^5^ using Jupyter Notebooks ^6^ and RStudio.^7^ A graphical user interface tool called the *Cohort Builder* allows researchers to identify cohorts of interest and create datasets extract data from the CDR using automatically generated SQL code.

While this setup has made the All of Us research program data widely available to researchers, a high technical burden for successful research remains. Studies beyond basic characterization often require intermediate to advanced programming skills in R, Python, and/or SQL and an understanding of the OMOP CDM, ultimately limiting the accessibility of the *All of Us* program. In our own experience using and mentoring students using the Researcher Workbench, this technical burden often slows research productivity and results in frustration. It can result in questionable research practices, and errors are often difficult to identify or fix. Similar experiences likely harm the research goals of the All of Us community and increase disparities in the successful use of the data between researchers with different training opportunities and backgrounds.

## OBJECTIVE

In response, we have developed the `allofus` R package to facilitate successful, reproducible use of the All of Us research program data. By increasing program accessibility and researcher competency for a diverse community of researchers, the `allofus` R package aims to build research capacity, ultimately providing value to the wider *All of Us* community. Our goals were to create a tool that would 1) make connecting to the database and managing files simple, 2) facilitate use of the popular `“`tidyverse” ecosystem of R packages^8^ on All of Us data, 3) help researchers more efficiently and accurately extract and synthesize survey data and EHR data, 4) increase the interoperability between fully-featured tools created by the OHDSI community and the Researcher Workbench. In addition, we have created a small but growing set of tutorials for researchers of all skills and backgrounds that demonstrate how to use the package.. In this article, we review the functionality of the `allofus` R package and demonstrate how it builds on existing Researcher Workbench resources.

## METHODS AND MATERIALS

### Package design

The `allofus` R package provides functions that address common challenges we encountered when working with All of Us Research Program Data. Table 1 describes these functions with respect to how they facilitate the use of All of Us data. The package is specifically designed to leverage the tidyverse ecosystem of R packages,^8^ including the dplyr package,^9^ to interact with the All of Us database. The general workflow for using the package functions is depicted in Figure 1. In this section, we briefly describe some of the key technical barriers to the successful use of the Researcher Workbench and All of Us Program database that are addressed by the `allofus` R package.

**Figure 1:**
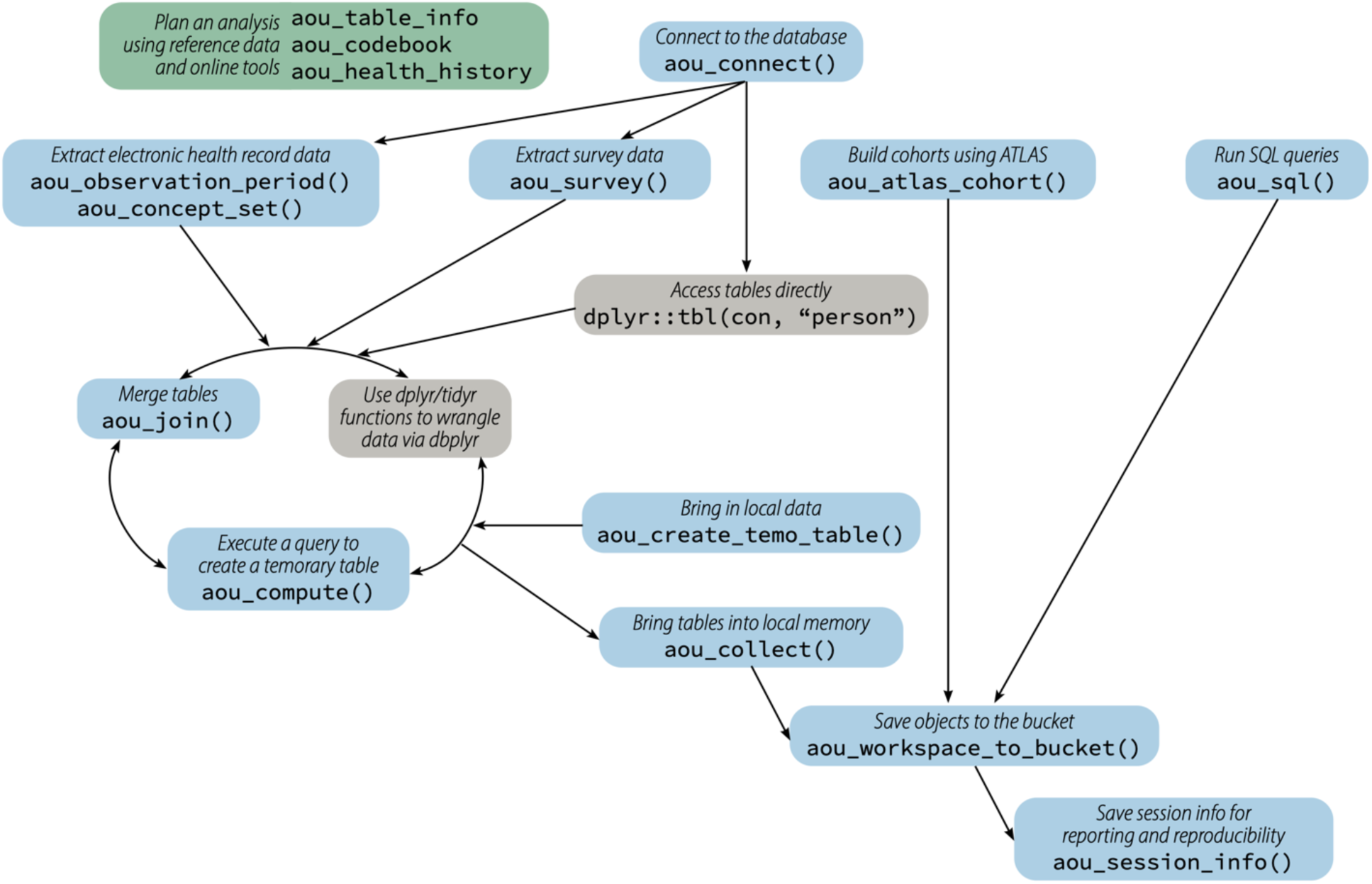
A simplified schematic of possible workflows on the All of Us Researcher Workbench using the `allofus` R package. Nodes in blue are package functions (a non-exhaustive list). The package is designed to integrate seamlessly with dbplyr (gray nodes), enabling dplyr functions on database backends. The package also provides several data tables, which are also available online, to help plan analyses.

**Table 1:**
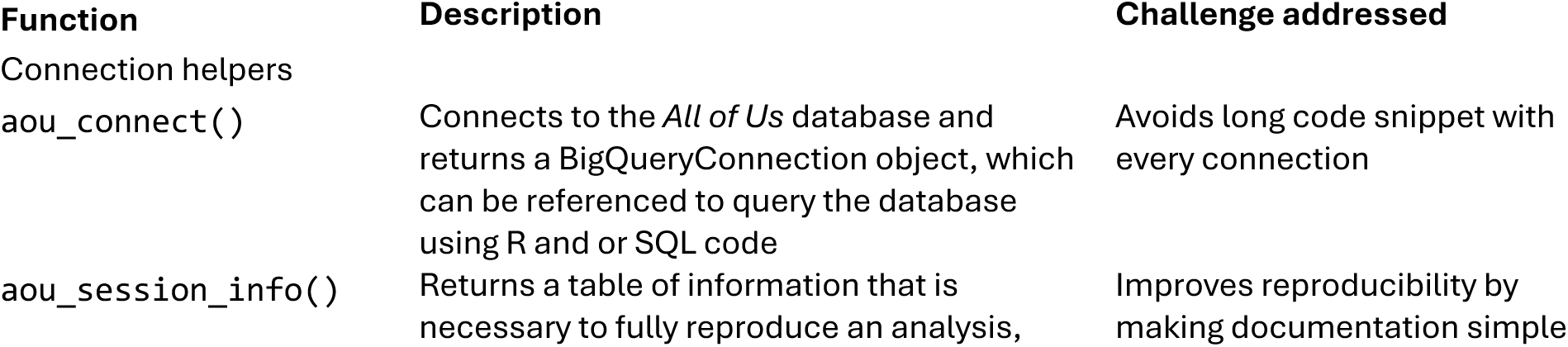
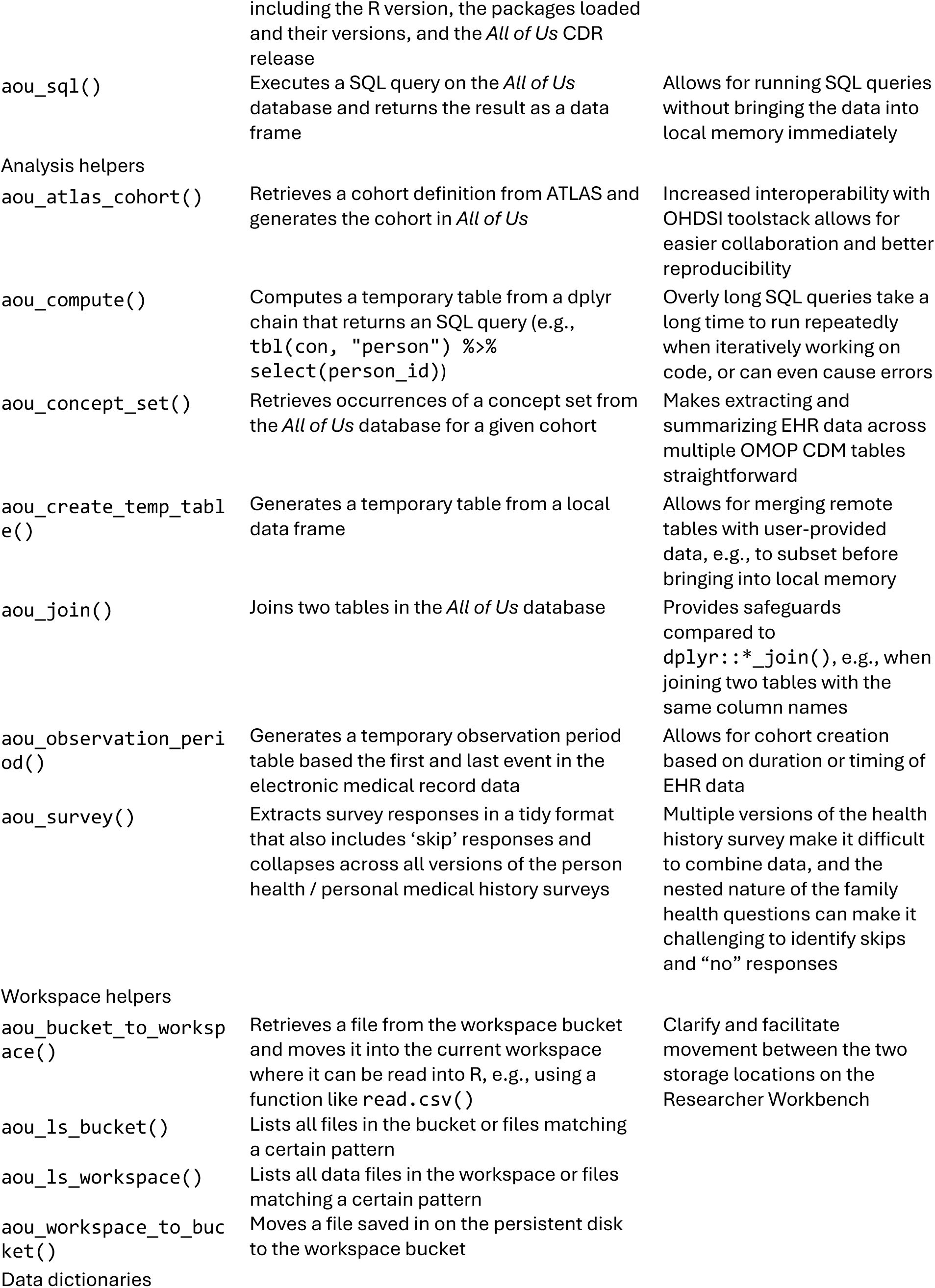
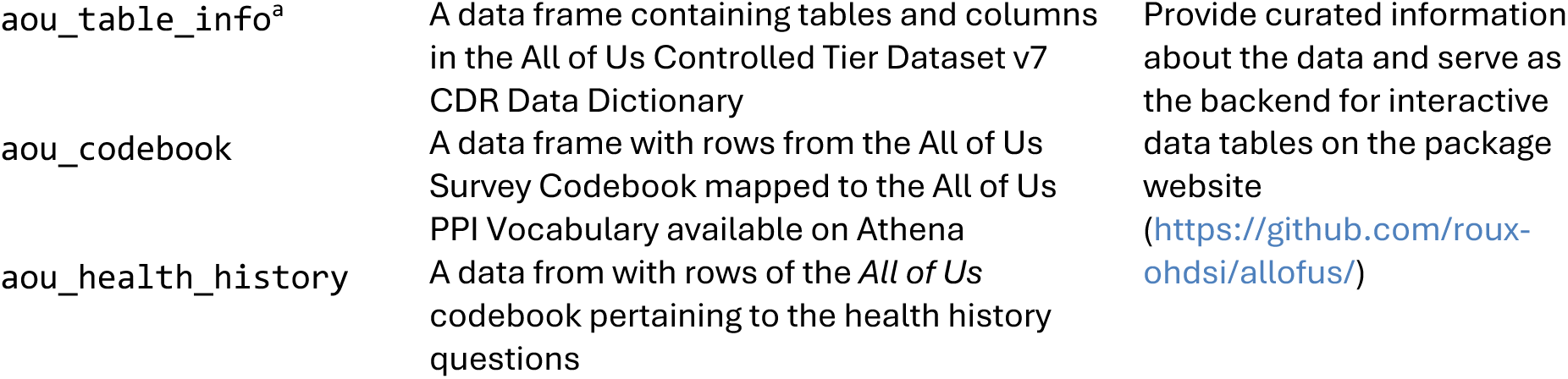
List of current functions and data available within the allofus R package.

#### Connecting to the database

Because All of Us data are stored centrally in a BigQuery cloud database,^10^ any analysis must connect to the database in some way. The All of Us Cohort Builder generates R or Python code that executes a SQL query and downloads the resulting table to the “workspace bucket” (a remote shared storage space for a collaborative workspace). Future analyses must move this file from the bucket to the “persistent disk” (a more temporary user-specific storage space) and load the data into local memory for analysis. Another option for interacting with BigQuery cloud database in R is via the `dbplyr`^11^ package, which allows researchers to use `dplyr`^9^ and other R functions directly on the database by translating them to SQL code. The `allofus` package takes advantage of this functionality with the `aou_connect()` function, which creates a BigQuery database connection using the `DBI` package^12^ that stays open as long as a notebook is in use. Many of the package functions work on the database via this connection but researchers can also directly use the database querying tools provided by `dbplyr` and `DBI`.

#### Writing and executing queries

A typical `dbplyr` query consists of a reference to one or more remote tables and a series of `dplyr` functions to manipulate data from the tables (e.g., `mutate()`, `filter()`, and `select()`) connected using a pipe operator. The SQL query it represents is only executed when the R object is printed or when the `collect()` function is used to bring the data into local memory. The `allofus` package provides several functions to facilitate this process within the constraints of the BigQuery database. When queries get too long or complex, they can result in SQL errors. The `aou_compute()` function forces early execution of the query, storing the results in a temporary table for additional manipulation. The `aou_collect()` function, which executes a query and downloads its result, is a wrapper for `dplyr::collect()`that deals explicitly with integer-valued columns that may be too large to be represented with base R’s integer class.

Along with the option to write code using `dplyr` syntax, users can also use the `aou_sql()` function easily send a SQL query to the database. References to database tables in the form of “{CDR}.table” will be automatically completed, and other objects or R code can be included within curly braces, allowing for parametrized queries. As with most of the package functions, the `collect` argument allows the user to choose to leave the query unexecuted on the database or downloaded into local memory.

Some queries rely on user-provided data. For example, filtering data for a large number of codes can be made more efficient by joining the data with a table of those codes. However, due to `bigrquery` limitations on temporary tables and lack of write access on the CDR, joining remote tables with local data is not straightforward. We wrote `aou_create_temporary_table()`, which creates a new temporary table from a local dataframe, to get around these limitations.

#### EHR and survey data tools

Survey and EHR data are standardized according to the OMOP CDM. This data model has been described in detail elsewhere;^3^ briefly, codes (e.g., ICD-10 codes) are mapped to standard “concepts”. Electronic health record research using OMOP often involves creating a list of the standardized codes (a “concept set”) that define a certain condition. Most concepts can be found by searching the public data browser, though some (e.g., those for devices) cannot. Alternatively, all concepts can be found using publicly available tools such as Athena (https://athena.ohdsi.org/). After a researcher has defined a concept set, the `aou_concept_set()` searches for instances of the selected codes across OMOP tables. Data can either be returned in full (i.e., all observations of the codes) or in summary (as a count of codes or indicator that at least 1 was present). Arguments to set the start and end date for each individual can restrict the extracted codes to a specific period.

The survey data are currently stored in two places in the database: the custom “ds_survey” table, which supports the Researcher Workbench GUI, and the OMOP “observation” table. Although the “ds_survey” table is more intuitively structured, it does not contain complete skip and non-response information for the health/medical history surveys. Therefore, our survey tools are based on the observation table, and are designed to address the complex nested nature of these data and encourage researchers to make explicit choices for dealing with missing data.

The `aou_survey()` function returns a table with a column for the response to each requested survey question and the response date. To find concepts representing questions of interest for the function, researchers can use two codebooks (`aou_codebook, aou_health_history`) in the allofus package (also available on the allofus package website) or the aforementioned publicly available tools. The codebooks are based on CDR v7.0 and contain information on the survey questions, the response options, and the codes used to represent those response options in the database.

#### Cohort creation

ATLAS^13^ is an open-source web app created by the OHDSI community that connects to an OMOP database and includes a vast array of tools for working with real-world data. Atlas’ concept set tools allow users to search for and get counts for concepts, much like the All of Us data browser. It also has functionality for creating complex cohort definitions (e.g., based on time-varying criteria), which are often shared among researchers to conduct network studies and for research consistency and reproducibility. Although ATLAS can’t be directly linked to the All of Us database, it can be used to define cohorts to be executed on All of Us data. Researchers without access to an ATLAS instance can use the free demo version (https://atlas-demo.ohdsi.org/) to define a cohort to be excuted using the `aou_atlas_cohort()` function on the Researcher Workbench.

Generating ATLAS cohorts requires an “observation period” table, which contains the dates under which individuals were “under observation”, i.e., when their health data would have been likely to be captured. The All of Us observation period table is not set up for this purpose, so we designed the `aou_observation_period()` function to create a table with the earliest and latest date each participant’s EHR data appears in the database. The function can also be used on its own to facilitate manual cohort construction. Because some EHR sites have contributed data from several decades ago, researchers might want to consider further constraining this table to reasonable date ranges of interest.

#### File management

The `allofus` package also includes helper functions to simplify file management. On the Researcher Workbench, files are stored in a Google Cloud bucket and must be moved via command line prompts. We designed a set of functions to list the files in the bucket or workspace associated with the current project and to copy files between the two locations, simplifying the Workbench code snippets. A comparison between the provided code snippets and the file management functions in the `allofus` R package is shown in Table 2.

**Table 2:** Comparison between provided command line code snippets and allofus R package functions for file management.

| Task | Provided code snippet | *allofus* function |
| --- | --- | --- |
| List files in the bucket | `# Get the bucket name my_bucket <- Sys.getenv('WORKSPACE_BUCKET') # List objects in the bucket system(paste0("gsutil ls -r ", my_bucket), intern=T)` | `aou_ls_bucket()` |
| Move a file from the bucket to workspace disk | `# replace 'test.csv' with the name of the file in your google bucket (don't delete the quotation marks) name_of_file_in_bucket <- 'test.csv' # Get the bucket name my_bucket <- Sys.getenv('WORKSPACE_BUCKET') # Copy the file from current workspace to the bucket system(paste0("gsutil cp ", my_bucket, "/data/", name_of_file_in_bucket, " ."), intern=T) # Load the file into a dataframe my_dataframe <- read_csv(name_of_file_in_bucket)` | `aou_bucket_to_workspace("test.csv")` |
| Write a file to disk and move it to the bucket | `# Replace df with THE NAME OF YOUR DATAFRAME my_dataframe <- df # Replace 'test.csv' with THE NAME of the file you're going to store in the bucket (don't delete the quotation marks) destination_filename <- 'test.csv' # store the dataframe in current workspace write_excel_csv(my_dataframe, destination_filename) # Get the bucket name my_bucket <- Sys.getenv('WORKSPACE_BUCKET') # Copy the file from current workspace to the bucket system(paste0("gsutil cp ./", destination_filename, " ", my_bucket, "/data/"), intern=T) # Check if file is in the bucket system(paste0("gsutil ls ", my_bucket, "/data/*.csv"), intern=T)` | `write.csv(df, "test.csv") aou_workspace_to_bucket(df, "test.csv")` |

### Package implementation and availability

We wrote the `allofus` R package for use on the Researcher Workbench (current R version 4.3.1) The primary dependencies are to the `bigrquery` R package,^14^ which facilitates connection to the Google BigQuery database, and to `dbplyr`, `dplyr` and other tidyverse packages. The package is designed specifically for use on the Researcher Workbench, includes unit tests for key functions, and is licensed under the MIT License.^15^ It is available for download on the R package repository CRAN (https://cran.r-project.org/web/packages/allofus/index.html), the source code for the package is available on GitHub (https://github.com/roux-ohdsi/allofus/), and a website with documentation, searchable reference tables, and tutorials is available at https://roux-ohdsi.github.io/allofus/.

## RESULTS

To illustrate the basic functions of the `allofus` R package, the following will describe the preparation of a simplified cohort of All of Us participants who do not have Type 2 Diabetes (T2DM), e.g., to look at risk factors for new diagnoses. We will demonstrate performing the data manipulation directly in the database rather than downloading and wrangling local data in the workspace. A Jupyter Notebook with the code is provided as Supplementary Material.

To define this cohort, we decide that we can be reasonably confident that participants have not been previously diagnosed with T2DM if they have at least 1 year of EHR data prior to joining All of Us without any codes related to T2DM. To identify those eligible, we identify each participant’s earliest survey date, subtracting 1 year, and joining this data to the table created by `aou_observation_period()`.

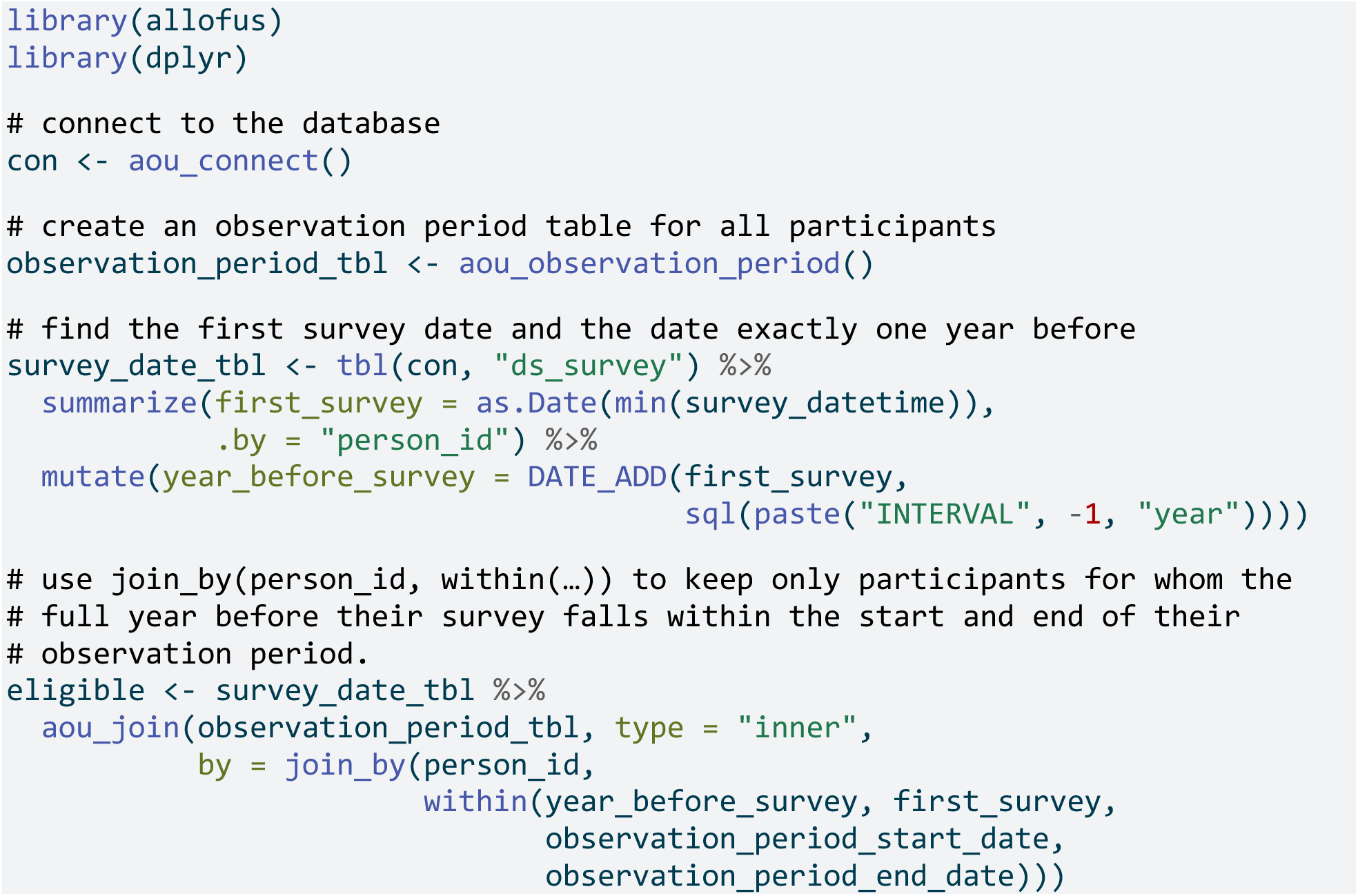

Next, we will consider someone to have a diagnosis of T2DM if they have at least 2 of: a diagnosis code for Type 2 diabetes, a prescription for metformin, a lab test for hemoglobin A1c measuring above 6.5%; or if they have a device for home blood glucose monitoring. In this toy example, we will use a non-exhaustive list of related concepts in Table 3, acknowledging that a real analysis should be more thorough and thoughtful about these choices.

**Table 3:**
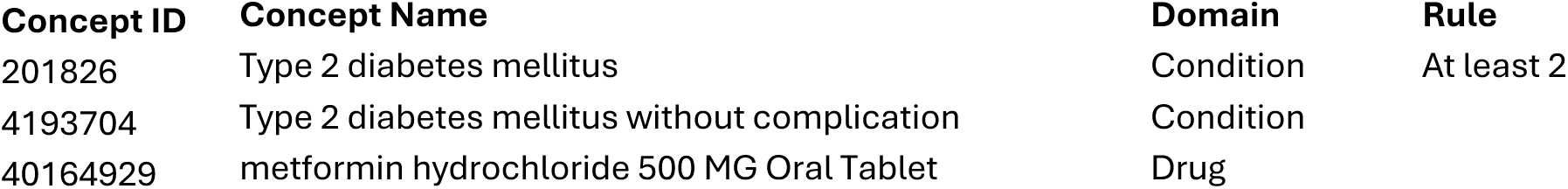
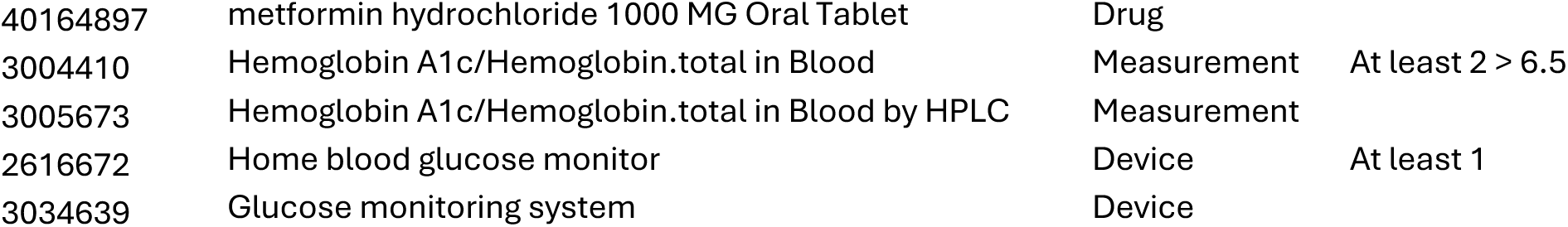
Example concept set with diabetes-related concepts from various domains, used to create a simplified cohort definition of All of Us participants with Type 2 Diabetes.

| Concept ID | Concept Name | Domain | Rule |
| --- | --- | --- | --- |
| 201826 | Type 2 diabetes mellitus | Condition | At least 2 |
| 4193704 | Type 2 diabetes mellitus without complication | Condition |  |
| 40164929 | metformin hydrochloride 500 MG Oral Tablet | Drug |  |
| 40164897 | metformin hydrochloride 1000 MG Oral Tablet | Drug |  |
| 3004410 | Hemoglobin A1c/Hemoglobin.total in Blood | Measurement | At least 2 > 6.5 |
| 3005673 | Hemoglobin A1c/Hemoglobin.total in Blood by HPLC | Measurement |  |
| 2616672 | Home blood glucose monitor | Device | At least 1 |
| 3034639 | Glucose monitoring system | Device |  |

We can extract the data using `aou_concept_set()`, combine it into a single table, and let it temporarily remain in the database so that we can perform more SQL manipulations. By default, `aou_concept_set()` will pull all matching codes from all OMOP tables across the entire timespan of data. However, to look for people with pre-existing T2DM, we need to specify `start_date` and `end_date` arguments to refer to the year before the initial All of Us survey. To identify people with sufficient evidence of T2DM, we use the function to request an indicator variable for the presence of at least 1 device code during that time period, a variable indicating whether a person has at least two of the condition or drug codes, and all of the related measurement data so that we can identify values above the minimum. Then we will combine these datasets into a single table containing participants with a previous T2Dm diagnosis (i.e., ineligible) and then remove them from our initial cohort.

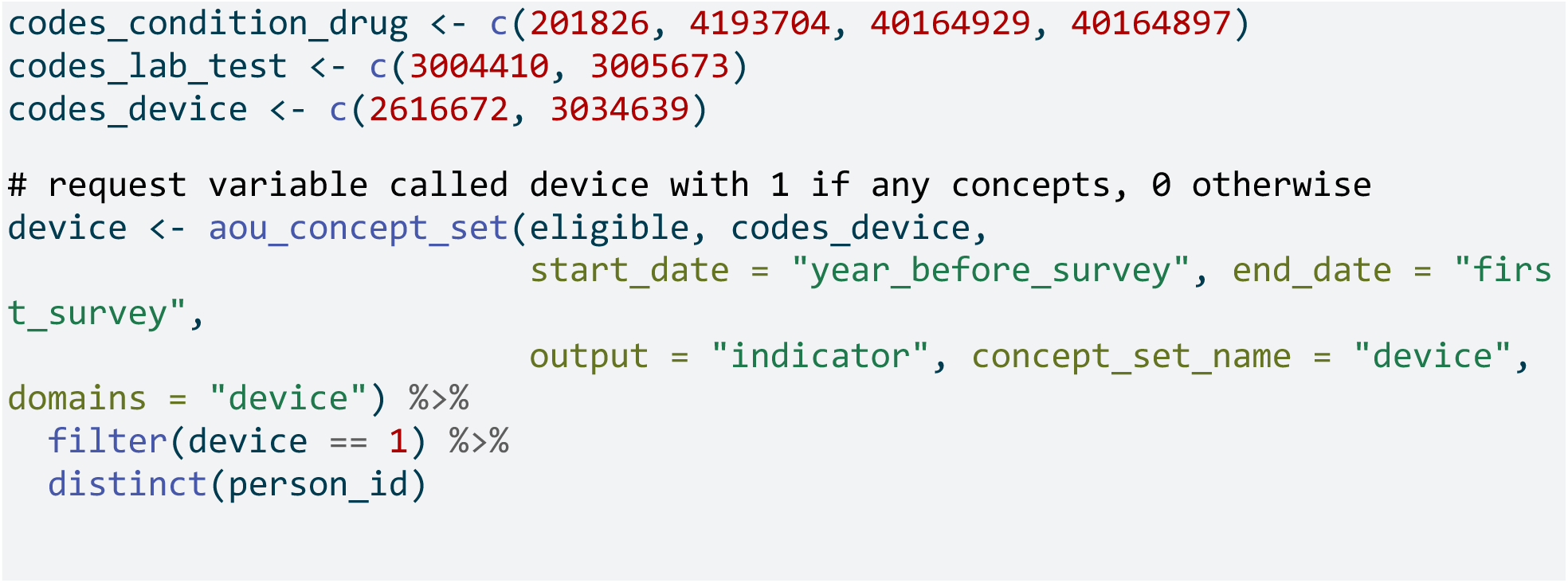
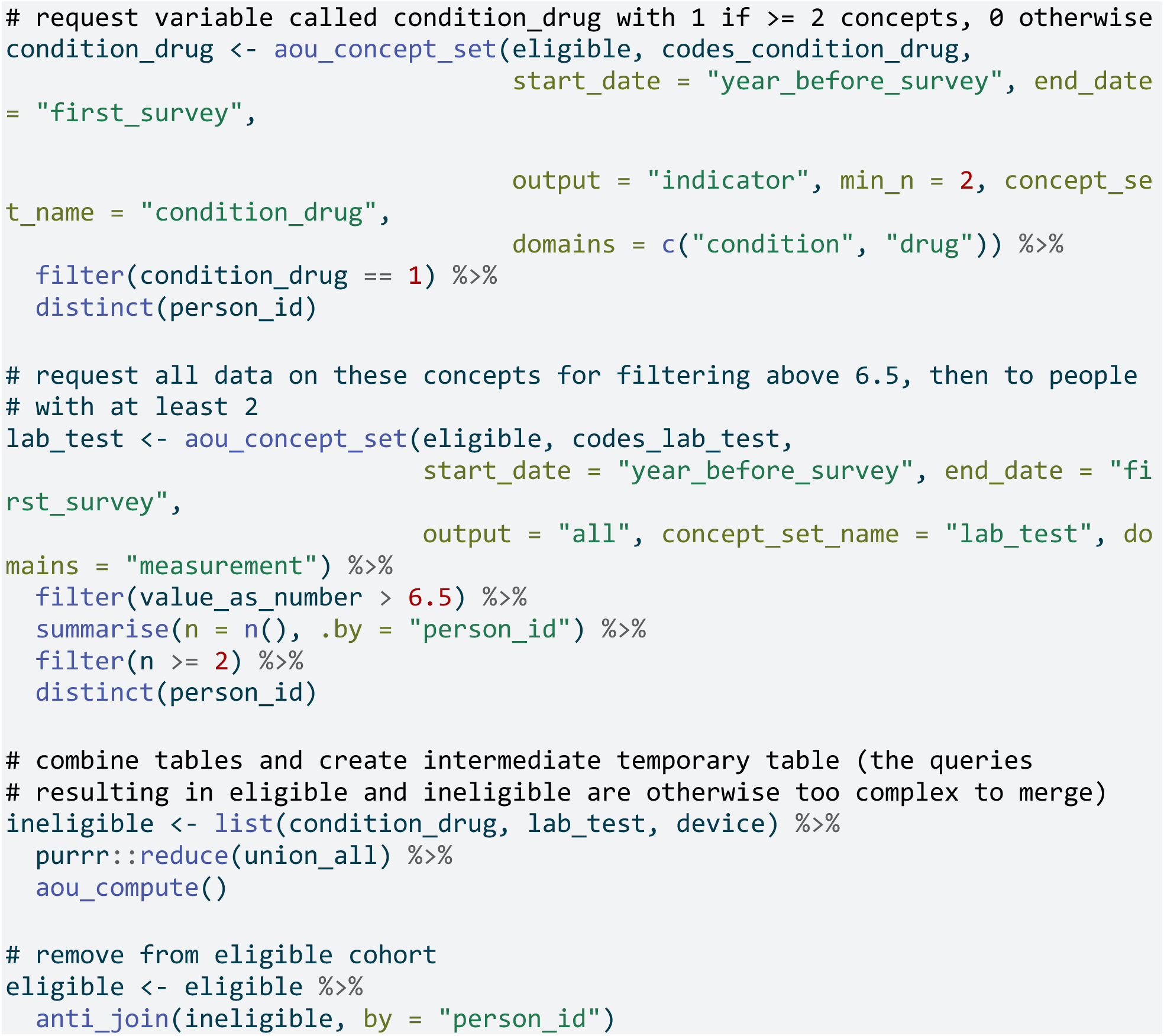

To illustrate how the `allofus` R package mitigates challenges related to the All of Us health-related surveys, we will also also confirm participants reported no prior T2DM diagnosis in the All of Us Personal and Family Health History survey or in the Personal Medical History, which replaced the former in 2021.^16^ Changes in the survey design make it difficult to correctly classify participants over time. This is reflected in the All of Us public data browser, which only shows a subset of participant responses for that answer–those reporting T2DM in the most recent, combined version of the survey.^17^

Concept ID 43529932 stores the response that indicates that a person self-reported T2DM (“Including yourself, who in your family has had type 2 diabetes? – Self”). In the initial Personal Medical History survey, the question that would elicit a respondent’s personal history of T2DM was “Has a doctor or health care provider ever told you that you have ?” with a list of endocrine conditions. The present Personal and Family Health History survey also asks about family medical history: “Have you or anyone in your family ever been diagnosed with the following hormone and endocrine conditions?” When a respondent selects a condition such as T2DM, they are then asked, “Including yourself, who in your family has had Type 2 diabetes?” with options including “self” and other family members. This multistage question makes it difficult to correctly combine data across both surveys and identify missing data and “no” responses. For example, the lack of a “self” response to the more recent question format could mean that the respondent didn’t answer any questions about endocrine conditions, that someone in their family does have T2DM but that individual does not, or that no one has any endocrine conditions in their family.

Along with “yes” (i.e., “self”) responses identified by the concept ID, the `aou_survey()` function handles the ambiguity of other and missing responses by classifying participants as “no” (assumed when someone answers that, for example, no one in their family has an endocrine condition, or they report that someone else but not themselves has type 2 diabetes or another endocrine condition), “don’t know”/“skip”/“prefer not to answer” (to either of the parent questions), or NA, which indicates that a person never completed a survey with that question. Other survey questions, e.g., about education and income (both from the Basics survey), are more straightforward.

To identify participants who self-reported T2DM (and extract information about education and income), we provide `aou_survey()`with our initial cohort of participants without an EHR indication of T2DM, concept IDs for self-report of T2DM, education, and income, and the desired names of the columns for each of these concept IDs. Joining back to our eligible table produces a final cohort of people who had one year of EHR data prior to the first survey, who had no indication of Type 2 diabetes in that period, and who took one of the two versions of the personal health history survey and reported that they did not have Type 2 diabetes.

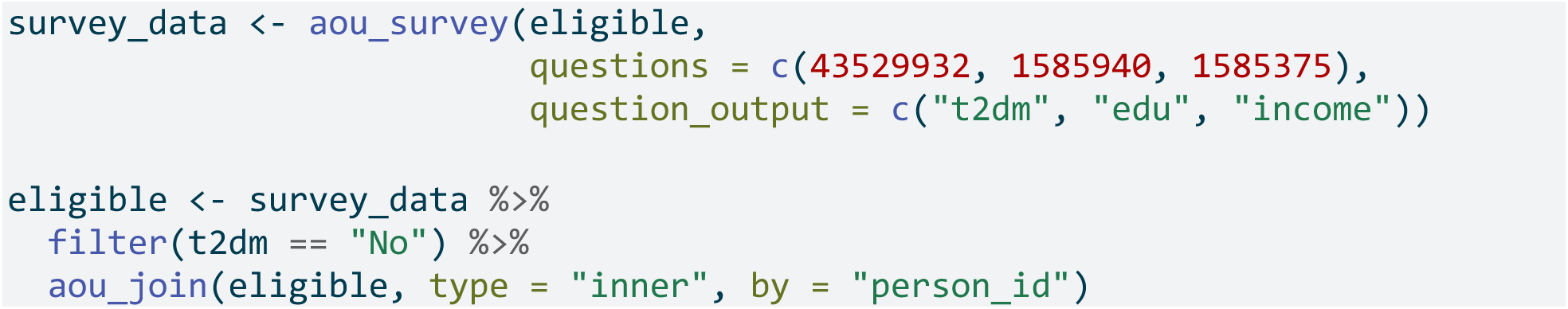

All data manipulation has taken place directly on the remote database, and this final table remains there. This minimizes the need to extract information on participants who have already been found ineligible in earlier steps. It also allows us to efficiently revise code if we make different decisions about defining the cohort. To save the data, make a table describing the cohort, or fit a regression model, we can bring it into memory using `aou_collect()`, which will convert integer-valued data to avoid errors in certain settings. This object is now a data frame that could be saved, e.g., as a .csv file. After saving, we can move it to the bucket for permanent storage and to share with collaborators with access to the project with `aou_workspace_to_bucket()`. Because the entire script was written in R with tidyverse-style code, rather than in SQL code generated by the cohort builder, sharing the script can ensure reproducibility and allow others to easily examine our analytic choices.

## DISCUSSION

In creating the `allofus` R package, we have attempted to reduce the technical burden and statistical programming knowledge required to conduct rigorous research on the All of Us research program database. The package stemmed from our experiences using the Researcher Workbench and mentoring students whose research projects use All of Us data. The functions in the package range from abstracting away bash commands and complex SQL code to extracting complex survey data, increasing compatibility with a key OHDSI tool, and allowing for easy linking of external data with the database. These tools enable R users to work more seamlessly with the All of Us program data directly and improve accessibility for beginner R users without threatening the rigor and validity of research. “Tidyverse-style” is often the preferred syntax for teaching new R users as it comprises a fully-featured data science ecosystem intentionally designed to ease the learning process with improved readability.^18^ By creating methods to make this ecosystem more compatible with the *All of Us* Researcher Workbench, data science tasks such as creating a cohort and extracting data from various database tables become more accessible to a wider range of researchers, who benefit from a consistent toolkit.^19^ We have published with the package a series of tutorials with commented code and explanations for extracting and wrangling the data in different ways. We plan to continue building these tutorials and invite other researchers to contribute their own via the package GitHub repository.

### Future directions

We have identified several key future directions for the package. First, we will continue to work on increasing interoperability with OHDSI tools such as ATLAS and Phenotype Library to allow researchers to leverage these tools in All of Us data. Additionally, while the current R package has focused primarily on the survey and EHR data, we aim to expand our attention to the other data sources: physical measurement, Fitbit, and genomics data.

Secondly, we aim to add methods that assess and mitigate biases resulting from missing data, selection bias, and lack of representativeness in *All of Us*. Although the All of Us mission is to ensure representation from groups historically underrepresented in research, the study has no sampling frame. Participation bias is thus difficult to assess but can threaten the validity of All of Us findings. In addition, there are varying degrees of missing data throughout the *All of Us* surveys (e.g., “skip” responses) that require thoughtful methods to address. Future package functionality will support addressing these biases.

## CONCLUSION

The `allofus` R package aims to return value to communities by training and building community research capacity. We hope that it will increase the accessibility of the All of Us research program for a diverse community of researchers as well as help generate rigorous research. Other researchers are encouraged to participate in this community project by suggesting and contributing extensions to the package.

## Data Availability

All of Us Research Program data are freely available to registered researchers.

https://roux-ohdsi.github.io/

https://github.com/roux-ohdsi/allofus

## Acknowledgments

We gratefully acknowledge *All of Us* participants for their contributions, without whom this research would not have been possible, as well as the National Institutes of Health’s *All of Us* Research Program for making available the data and research platform on which this work was conducted.

We also wish to thank Chloe Bennett, Narcissa Plummer, and Chelsea Wong for helping test the package’s functionality.

